# Estimating COVID-19 Cumulative Incidence from Seroprevalence Surveys accounting for Time-Varying Seroreversion: A Fully Bayesian Methodology

**DOI:** 10.64898/2026.06.09.26355264

**Authors:** Nana Owusu-Boaitey, Mark J. Meyer, Daniel Herrera-Esposito, Lucas Böttcher, Maria Lukz, Sydney Cook, Michael A. Stoto, John D. Kraemer

## Abstract

Seroprevalence surveys reveal the extent of humoral immunity against pathogens such as severe acute respiratory syndrome coronavirus 2 (SARS-CoV-2), and under some circumstances represent cumulative incidence of prior infection. However, antibody waning–or seroreversion– biases these estimates by reducing assay sensitivity in a time-varying manner. Because assay sensitivity decays over time, naively using serosurveys can substantially bias estimates of SARS-CoV-2 cumulative incidence and fatality rates. The Bayesian assay-specific, time-varying sensitivity adjustment developed in this paper can reliably correct for this bias and account for the delay between infection and serosurvey. In seroprevalence studies conducted in the United States in 2020, adjusting for time-varying sensitivity increased cumulative incidence by up to 1.4-fold, with an adjustment of 1.08 for a national study. Our estimates contrast with a previously published 2-fold adjustment that did not account for assay design. This suggests that previous analyses overestimated cumulative incidence by applying seroreversion corrections that did not account for assay-specific effects, or underestimated cumulative incidence by not applying seroreversion corrections. These biases imply fatality rate underestimation and overestimation, respectively. Our model provides a framework for design-specific time-varying sensitivity corrections in seroprevalence surveys for other pathogens.

**Topic:** diagnostic sensitivity, seroprevalence, SARS-CoV-2, COVID-19

## Introduction

Understanding the extent to which transmission of an infection has occurred is of critical public-health importance. Cumulative incidence—estimated by proportion of individuals with serological evidence of a prior infection with a pathogen—can inform policy by indicating the number of individuals with acquired humoral immunity as well as how the risk of infection and acquisition of immunity varies over time and among population subgroups. It also allows estimation of other metrics, such as the infection fatality rate (IFR) and infection hospitalization rate (IHR) that are useful measures of disease severity.

However, estimating cumulative infection rates can be challenging. For example, during the COVID-19 pandemic, observed case counts—based on cases reported by clinicians and labs— were the primary method for tracking epidemiology. But these counts were severely biased both by asymptomatic and minimally symptomatic disease leading to a lack of diagnosis and limited testing capacity. Studies suggest that as few as 10% of cases were detected during the first 4 months [1–3].

One method for estimating cumulative incidence is to use the prevalence of antibodies against the virus SARS-CoV-2. Antibodies specific to this virus reliably increase following infection, allowing prevalence of antibodies, or seroprevalence, to detect SARS-CoV-2 infections missed by testing [4,5]. Seroprevalence surveys of a representative sample of the population can provide useful estimates of the cumulative incidence of prior infection [6–8].

However, for seroprevalence to represent cumulative incidence, several sources of bias must be addressed. First, the assays used to estimate seroprevalence are imperfectly sensitive and specific. Thus, to obtain the true seroprevalence requires adjusting observed seroprevalence to account for the expected amount of both false positives and false negatives. There are several approaches for doing this including the fully Bayesian approach developed by Meyer et al. [8], which also provides valid uncertainty estimates.

A second problem is that antibody concentration wanes over time, which creates further bias when inferring cumulative incidence from seroprevalence [9,10]. Antibody titers can decrease below a particular serological assay’s limit of detection. This seroreversion reduces assay sensitivity with time following infection [8,10–12], which may cause observed seroprevalence to decrease with time in longitudinal testing of populations [13]. Thus, it is not sufficient to adjust seroprevalence estimates for sensitivity at a single baseline time point, but rather the adjustment must account for how sensitivity declines over time. If this is not done, seroreversion will cause seroprevalence to meaningfully underestimate post-infection immunity and cumulative incidence [7,14]. Adjusting seroprevalence using time-varying sensitivity estimates can correct for seroreversion and better approximate cumulative incidence [8,10,15].

Seroreversion adjustment requires assumptions regarding pandemic severity and immunity that can impact the estimates. Takahashi et al. [14], for example, argued seroreversion biased estimates more strongly than suggested by a prior study [6]. Buss et al. [7] inferred substantial seroreversion among previously infected residents of Manaus, Brazil, such that post-infection immunity missed by serology might be sufficient for herd immunity and prevention of further waves of infection. Later analyses implied less seroreversion, consistent with a further wave that occurred in Manaus [11,14,16,17]. Assay-specific differences in time-varying sensitivity could also explain why different assays administered to the same population yield divergent seroprevalence estimates [15].

Several different approaches to seroreversion correction have been used, though failing to account for seroreversion remains a persistent problem [18]. Some analyses applied the same correction across assays [1,6,19,20]. COVIDVu seroprevalence studies, for example, applied the anti-spike indirect ELISA adjustments of Shioda et al.[21] to an anti-nucleocapsid direct ELISA[1,22,23]. Other studies used assay-specific adjustments for a small number of assays [5,15]. Seroprevalence surveys also detected seroreversion using individuals in their sample with a previously diagnosed SARS-CoV-2 infection, or prior-positives, who then tested seropositive [2,24,25]. Levin et al. [5] combined data on prior-positives across multiple study populations to calculate time-varying sensitivity for one assay. Barber et al. [26] instead used prior-positive data from several studies to estimate time-varying sensitivity for eight assays. They extended their model to out-of-sample assays based on the isotype of, and SARS-CoV-2 antigen targeted by, antibodies detected by the assays. Other assay characteristics were not included.

Because some assays exhibit more time-varying sensitivity than others, seroreversion adjustments must take the assay into account. We illustrate the potential for this issue in Figure 1. The unprecedented number of seroprevalence surveys performed during the SARS-CoV-2 pandemic permitted better quantification of the contribution of assay design to time-varying sensitivity. Owusu-Boaitey et al. [12] used data on prior-positives to calculate assay-specific time-varying sensitivity for fifty assays, while also quantifying the contribution of assay design and antigen target to time-varying sensitivity. This quantification yielded design-specific and antigen-specific predictions consistent with time-varying sensitivity for two out-of-sample assays employed in seroprevalence studies [27,28]. The analysis was restricted to before the onset of widespread SARS-CoV-2 vaccination and thus reflects seroprevalence induced by infection instead of population immunity from vaccination.

**Figure 1.**
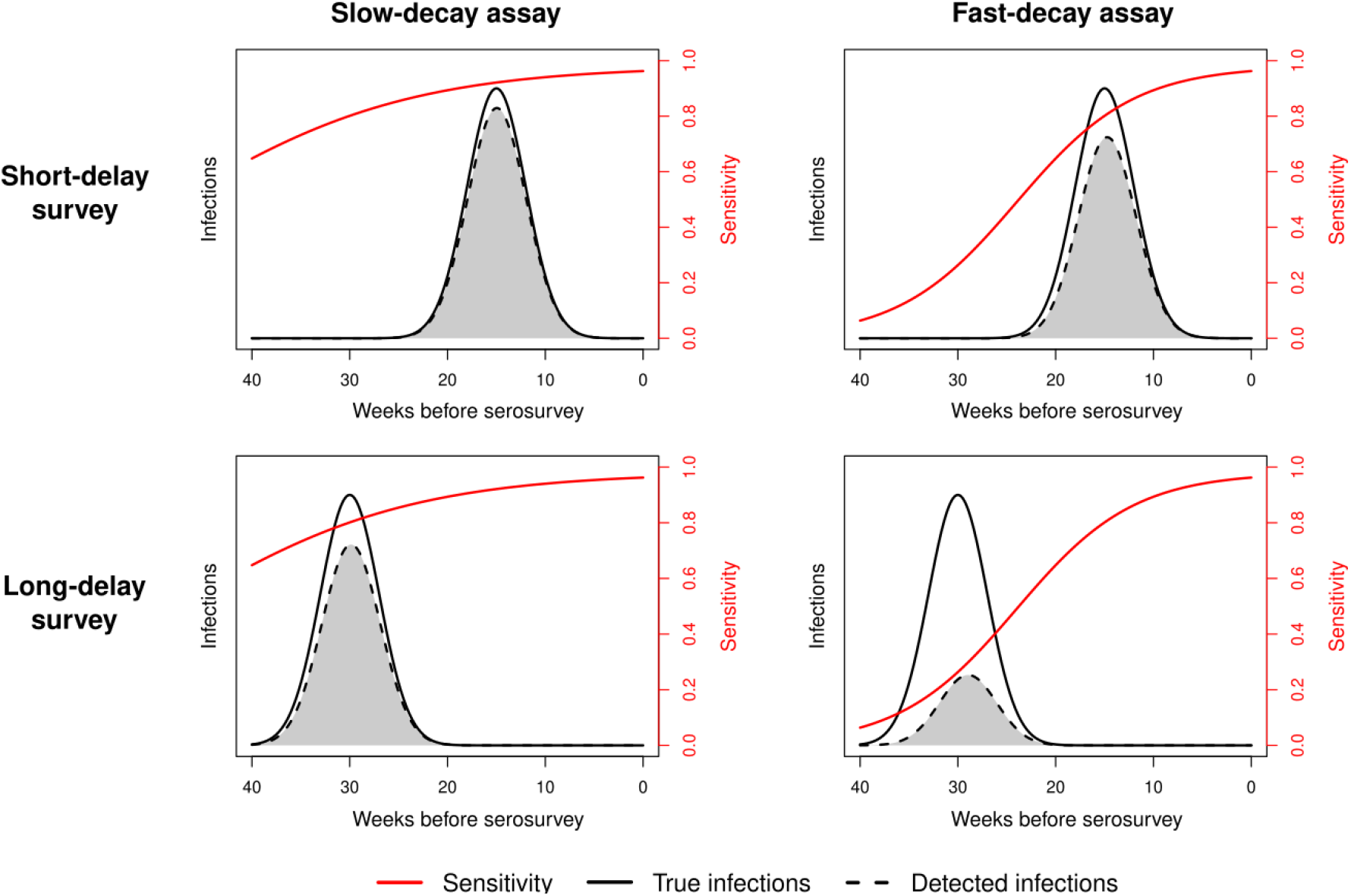
Example of seroreversion-induced biases. Each panel shows different sensitivity and infection dynamics prior to a serosurvey study. The red line indicates the time-dependent sensitivity of an assay for detecting infections that occurred before the serosurvey. In the left column, the sensitivity curve decays slowly; in the right column, it decays more rapidly. The solid black line represents the true number of infections of an epidemic wave (same number of infections for each panel). In the top row, there is a short delay between the epidemic wave and the serosurvey (infections further to the right); in the bottom row the delay is longer (infections further to the left). Lastly, the dashed black line indicates the number of infections detected by each serosurvey. It is obtained by multiplying the assay’s sensitivity by the true infections at each time point, neglecting false positives for expository simplicity. The proportion of detected infections depends strongly on the interaction between the assay-specific sensitivity decay and the serosurvey timing.

The present work develops a Bayesian approach for these assay-specific time-varying sensitivity estimates [12] to infer seroreversion-corrected cumulative incidence from seroprevalence surveys conducted in the United States before the onset of SARS-CoV-2 vaccination. This approach mitigates the impact of time-varying sensitivity differences across assays and accounts for the delay between infection and serosurvey. It also yields more realistic cumulative incidence estimates that incorporate uncertainty from time-varying sensitivity and from the number of samples used to estimate assay specificity [29]. In order to test the method and see how much difference the phenomenon can make, we applied this model to seroprevalence studies to obtain more accurate estimates of cumulative incidence of SARS-CoV-2 infection in 2020. Our estimates of the contribution of assay design to time-varying sensitivity and cumulative incidence estimates could be applied to assays for other pathogens.

## Methods

### Data

We constructed a model to estimate COVID-19 cumulative incidence from population-representative seroprevalence surveys using three pieces of information for each population: observed seroprevalence data, information on how the distribution of time elapsed between infection and the seroprevalence surveys (here using the observed pattern of excess mortality), and information on how assays’ sensitivity declines over time after infection.

This study estimates cumulative incidence using previously published seroprevalence surveys that employed probability sampling of the general population. Web Appendix S1 includes selection criteria for the seroprevalence surveys and the assessed seroprevalence studies. All included studies were performed in 2020, before the onset of widespread SARS-CoV-2 vaccination. We initially included only US studies to have relative consistency in data quality. However, we subsequently added a series of UK studies as a sensitivity analysis in order to capture more variation in assays’ risk of seroreversion and greater variation in the length of time after infection. For each study we recorded the region sampled, sampling start and end date, median date, serological assay(s) used for testing, number of individuals tested, and reported seroprevalence without adjustments for sensitivity and specificity. Population-weighted seroprevalence was used if provided by the seroprevalence study; otherwise, unweighted seroprevalence was recorded. We also recorded time-invariant assay sensitivity and specificity provided by the assay manufacturer or from another source, consistent with a prior publication [1].

Weekly excess mortality data for each region sampled was collected from a publicly available dataset [2,30]. Excess deaths were tabulated by date of death from the week of March 1, 2020, to three weeks past the median date of serological sampling. Three weeks was selected based on a three-week lag between infection to death, assuming one week lags between infection, symptom onset, seroconversion, and death [3–6,31,32]. While other sources of information about the timing of infections in a population—such as observed hospitalizations—could also be used, we based our estimates on excess mortality because it is expected to be least sensitive to limited testing capabilities or hospital capacity.

Assay-specific sensitivity estimates for each month following infection were taken from Owusu-Boaitey et al. [12]. These estimates were derived from the proportion of individuals with a previously diagnosed SARS-CoV-2 infection who tested seropositive in seroprevalence studies performed on the general population. This proportion was used as an estimate of assay sensitivity and data on this proportion was pooled together for studies that used the same assay. Reported median time between infection diagnosis and serological testing was taken from the seroprevalence studies. In cases where studies did not report this median time, it was inferred from reported case curves. From this data Owusu-Boaitey et al. inferred assay-specific monthly estimates of sensitivity up to 14 months following infection. Assays were also pooled by assay design, antibody isotype and antigen target of antibodies to quantify the contribution of these factors to time-varying sensitivity. Based on these estimates [12], we classified lateral flow assays and anti-nucleocapsid indirect ELISAs as high risk from seroreversion, while other assay types were classified as low risk. Assay-specific monthly estimates of sensitivity were used for each assay, except for the ‘University of Oxford assay for which assay-specific estimates were not provided in Owusu-Boaitey et al. For this assay we instead used anti-spike indirect ELISA monthly estimates from Owusu-Boaitey et al. to match the antigen target and assay design for the University of Oxford assay.

### Statistical analysis

The standard correction for sensitivity and specificity (known as the Rogen–Gladen formula [33]) has the form

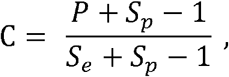

Where *C* denotes the cumulative incidence, *P* denotes the proportion reactive, *S*_*p*_ denotes the specificity, and *S*_*e*_ is the sensitivity. Each of these quantities is a proportion, although under the standard correction *C* can be below 0 or above 1. Any model employing this needs to take that into account. An alternative representation re-expresses this quantity as

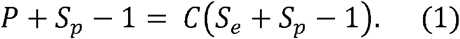

Let *Y*= *P* + *Sp* −1. By this definition, *Y* is no longer bounded by below by 0 but by −1 and is still bounded above by 1. Further, we assumeis *Y* measured with error since it depends on two unknown quantities that both must be estimated. Let *C* = *D β* where *D* is an *N × k* design matrix and *β* is a *k ×*1 vector of cumulative incidences to estimate. The quantity *k* is the number of seroprevalences to be estimated. For *P* and *D*, we use binary data representations. Thus, *P* is a *N ×* 1 vector of 0s and 1s with 1 corresponding to positive reactivity and 0 to negative. For a single estimated incidence, the data for *D* can simply be a vector of 1s with length equal to *P*. If estimating multiple incidences, clustered by region or surveys using the same assay for example, *D* becomes an appropriately structured matrix of indicator variables.

The right-hand side of the Equation (1) is then *C* (*S*_*e*_+*S*_*p*_ −1) *D β C* (*S*_*e*_+*S*_*p*_ −1). Since *C* (*S*_*e*_+*S*_*p*_ − 1) is a scalar, this term can be rewritten as *C* (*S*_*e*_+*S*_*p*_ −1) *Dβ* when estimating a single incidence. Thus, we consider the following linear model to estimate the unknown parameter *β*:

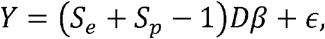

where *ϵ* is a mean zero Gaussian error term with variance 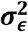. This model can be simplified by defining *X* = (*S*_*e*_+*S*_*p*_ −1) *D*, giving the linear regression form

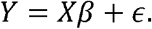

When only one seroprevalence is estimated (*k* =1), this is just a simple linear model.

The parameter *β* only has non-zero support on the unit interval and must be bounded. To enforce this constraint, we employ a fully Bayesian approach and impose a truncated normal prior on the parameter with truncation to the interval {0,1} and variance 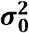 which we fix at a large value, e.g. 100 000. We place an inverse gamma prior on 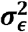 with parameters *A*_*∈*_ and *B*_*∈*_, both set to something small, e.g. 0.25.

Sensitivity and specificity can be fixed at specific values or their posteriors can be estimated first and used when sampling from the posterior of *β*. The latter approach naturally incorporates the variability of the sensitivity and specificity estimates into the estimation of the cumulative incidence. To estimate specificity,*S*_*p*_, we select a Beta conjugate prior for *S*_*p*_ with prior parameters chosen to be small (0.1), thus limiting their influence. We then draw from its posterior distribution where the data for the likelihood is determined by the test’s reported characteristics; specifically the number of false positives (*FP*) and the number of true negatives (*TN*), see [12]. In context, the prior parameters are equivalent to adding 0.1 true negatives and 0.1 false positives.

We estimate the posterior for *S*_*e*_ by first generating a posterior distribution for the seroreversion curve [12], with estimation occurring on a weekly time scale. Drawing on excess mortality data to estimate the average time since infection for respondents to seroprevalence surveys, we allow S_*e*_ to vary over time. This approach uses a hierarchical logistic model for estimation with predicted values representing weekly seroreversion curves. We then weight the predicted posterior by the proportion of weekly excess mortality over the desired interval as an indication of the estimated time lag between infection and the survey date. This creates an excess mortality adjusted seroreversion curve and corresponding distribution. After averaging across time, we obtain a posterior distribution of excess mortality adjusted seroreversion which we use for the sensitivity estimate in *X*, i.e. *S*_*e*_ . The values from the posterior distribution of *S*_*e*_ and *S*_*p*_ are fed into the Gibbs sampler for the posterior of *β* in a one-to-one fashion. That is, for each pair of samples from the joint posterior of {*S*_*p*_, *S*_*e*_}, we obtain one draw from the posterior of *β*. The estimation procedure is described in Algorithm 1. Exploratory empirical studies suggest this approach performs well in recovering the “true” cumulative incidence.

#### Algorithm 1

Procedure for estimating cumulative incidence, adjusted for excess mortality and seroreversion. The subscript {0,1} indicates truncation to the unit interval, *I* _*k*×*k*_ is a *k* ×*k* identity matrix, *InvGamma* denotes the inverse gamma distribution, and *n* is the sample size. The notation^(*m*)^ indicates the *m* th draw from the posterior while ^(m−1)^ indicates the (m −1) st draw.

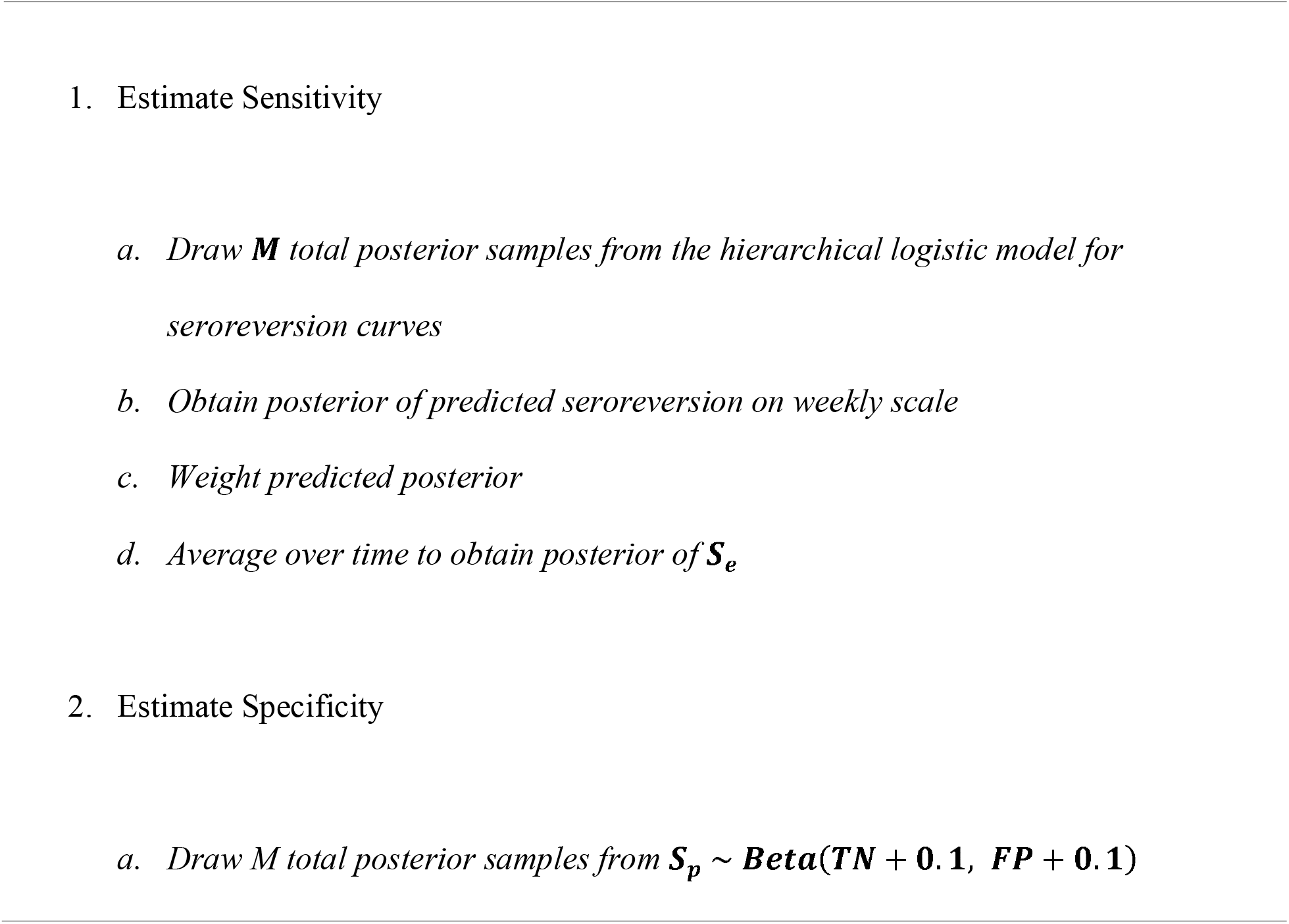
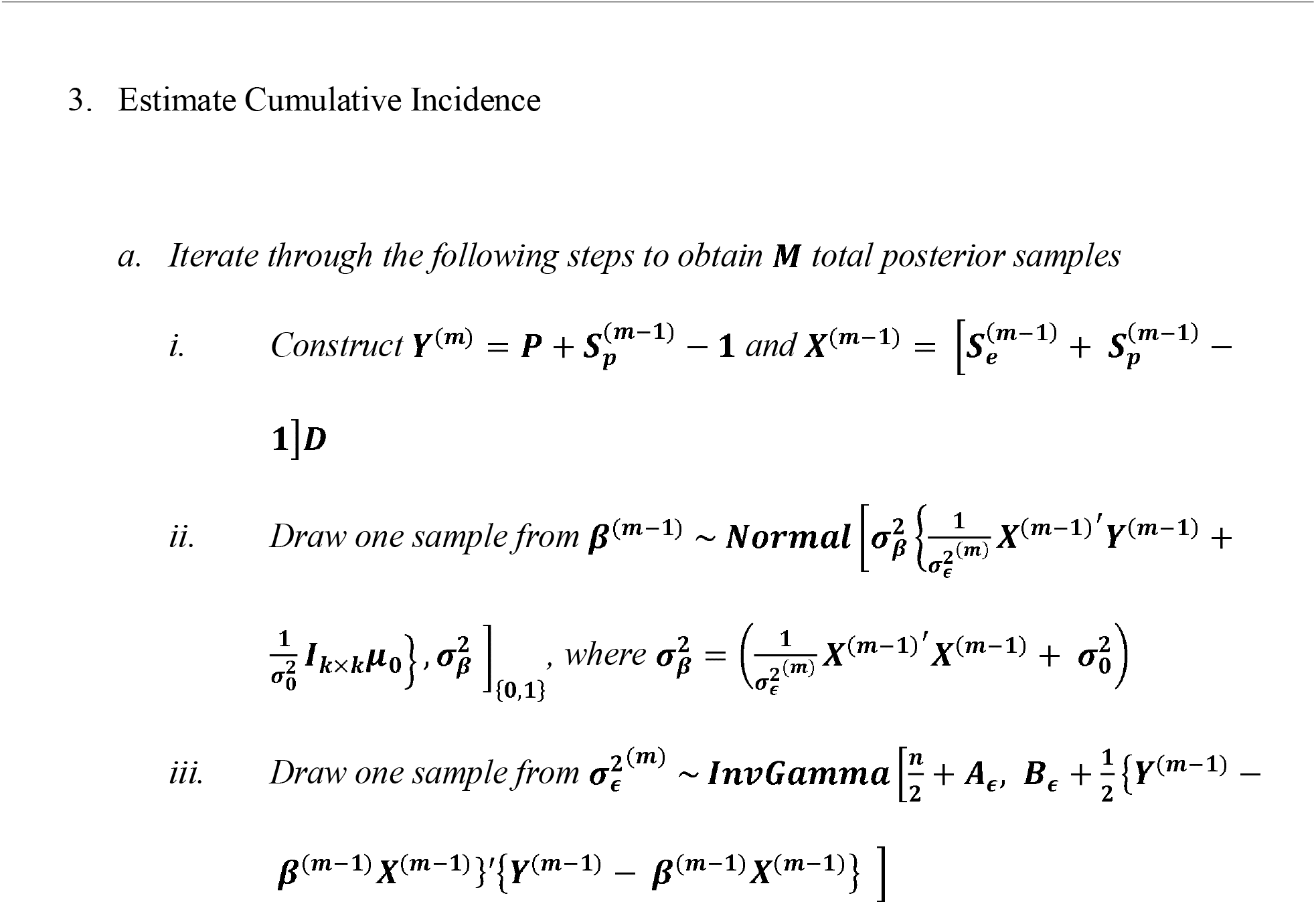

To represent the impact of the assay-specific time-varying adjustment, we report the ratio of adjusted seroprevalence estimates to the naive or unadjusted estimate from the original studies, defined as ratio

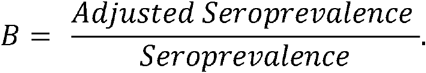

We calculate this metric in two ways. First, we adjust seroprevalence for test-specific sensitivity and specificity at the time of test, i.e. a time-invariant adjustment. This estimation is done using the method described in Algorithm 1 but setting *S*_*e*_ equal to the time 0 estimate of seroreversion. Second, we implement Algorithm 1 with a time-varying *S*_*e*_ calculation as described above. This gives us both time invariant and time-varying adjusted seroprevalence. We construct the posterior distribution of *B* under both models.

## Results

We have calculated adjustment factors for 15 surveys conducted in 11 jurisdictions in the United States (Table 1). We also report the posterior probability that the adjustment factor is greater than 1; that is, the probability that the adjusted seroprevalence is larger than the reported uncorrected seroprevalence. Estimated adjustment factors for the time-invariant estimates range from 0.938 (95% CrI: 0.428, 1.407) in Puerto Rico to 1.214 in Georgia (95% CrI: 0.954, 1.779). For all surveys except the one conducted in Puerto Rico, it is more likely than not that applying the time-invariant correction results in higher estimated seroprevalence than the reported uncorrected estimate.

**Table 1:** Posterior estimates of the adjustment factor, □, comparing reported seroprevalence to adjusted under both time invariant and time-varying adjustments. Estimates represent posterior medians and are accompanied by 95% credible intervals.

| Jurisdiction | Average | Risk from | Time-invariant/Reported | Time-varying/Reported |
| --- | --- | --- | --- | --- |

| | Interval from Infection (weeks) | seroreversion | Est. (95% CrI) | $P(B > 1)$ | Est. (95% CrI) | $P(B > 1)$ |
| --- | --- | --- | --- | --- | --- | --- |
| United States | 18 | Low | 1.207<br>(0.994, 1.763) | 0.971 | 1.289<br>(1.009, 2.217) | 0.980 |
| Arkansas |  |  |  |  |  |  |
| Phase 2 | 4 | Low | 1.051<br>(0.409, 1.789) | 0.560 | 1.066<br>(0.419, 1.805) | 0.576 |
| Phase 5 | 9 | Low | 1.111<br>(0.756, 1.575) | 0.729 | 1.129<br>(0.778, 1.658) | 0.756 |
| Phase 7 | 11 | Low | 1.156<br>(0.826, 1.632) | 0.811 | 1.174<br>(0.835, 1.706) | 0.831 |
| California | 16 | Low | 1.208<br>(0.824, 1.867) | 0.843 | 1.280<br>(0.855, 2.154) | 0.895 |
| Connecticut | 12 | Low | 1.117<br>(0.661, 1.569) | 0.692 | 1.238<br>(0.732, 1.825) | 0.813 |
| Georgia | 17 | Low | 1.214<br>(0.954, 1.779) | 0.942 | 1.287<br>(0.976, 2.168) | 0.959 |
| New York City | 18 | Low | 1.133<br>(0.927, 1.359) | 0.896 | 1.167<br>(0.956, 1.398) | 0.937 |
| Ohio | 9 | High | 1.049<br>(0.163, 1.996) | 0.541 | 1.225<br>(0.182, 2.314) | 0.657 |
| Puerto Rico | 17 | High | 0.938<br>(0.428, 1.407) | 0.398 | 1.319<br>(0.585, 2.272) | 0.802 |
| Rhode Island | 4 | High | 1.088<br>(0.639, 1.496) | 0.661 | 1.142<br>(0.679, 1.558) | 0.735 |
| Indiana |  |  |  |  |  |  |
| Phase 1 | 3 | High | 1.069<br>(0.414, 1.438) | 0.643 | 1.110<br>(0.416, 1.493) | 0.700 |
| Phase 2 | 7 | High | 1.073<br>(0.579, 1.436) | 0.646 | 1.175<br>(0.643, 1.574) | 0.795 |
| Wisconsin |  |  |  |  |  |  |
| Phase 1 | 10 | High | 1.075<br>(0.436, 1.643) | 0.602 | 1.296<br>(0.514, 1.993) | 0.791 |
| Phase 2 | 11 | High | 1.102<br>(0.836, 1.368) | 0.777 | 1.364<br>(1.030, 1.712) | 0.983 |
| Arkansas data used the Beckman ACCESS assay. California and Georgia used BioRad Platelia. Connecticut used Vitros Ortho, S CoV2 IgG. New York City used DiaSorin. Ohio, Rhode Island, Indiana, and Wisconsin used Abbott Architect. Puerto Rico used Abbott Alinity i. |  |  |  |  |  |  |

Adjustment factors that compare time-varying estimates to the reported unadjusted estimates deviate farther from equivalence and for all surveys it is more likely than not that the correction results in higher estimated seroprevalence. The smallest adjustment factor was the 1.066 observed in Arkansas at an average of four weeks post-infection (95% CrI: 0.419, 1.805). The largest was Wisconsin at an average of 11 weeks post-infection: 1.364 (95% CrI: 1.030, 1.712). Figure 2 suggests that the largest differences between time-invariant and time-varying adjustment factors occur for those seroprevalence surveys that were conducted later in the location’s epidemic and thus had a longer average time from infection to survey, as well as for surveys that used assays with a greater susceptibility to seroreversion.

**Figure 2:**
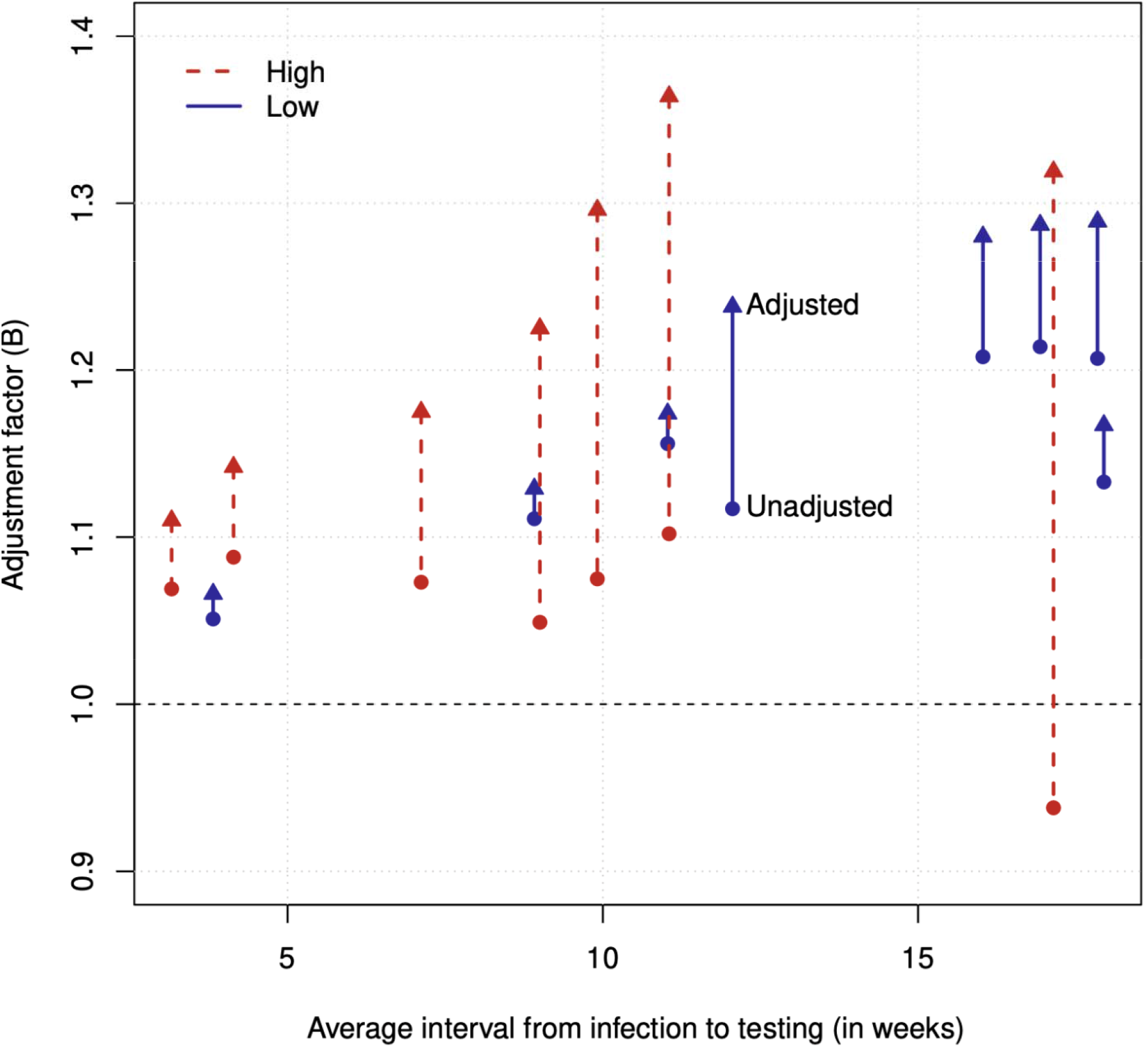
Adjustment factor when calculated using the time-invariant sensitivity (circles) versus calculated using time-varying sensitivity (triangles). Solid blue indicates low test-specific seroreversion risk while dashed red indicates high seroreversion risk.

When considering only surveys that used an assay with a high susceptibility to seroreversion, the degree of deviation between the time-invariant and time-varying adjustment factors increased monotonically with time. For example, the first Indiana sample (with an average of only 3 weeks passing before the survey) had a time-invariant adjustment factor of 1.069 (95% CrI: 0.414, 1.438) compared to a time-varying adjustment factor of 1.110 (95% CrI: 0.416, 1.493). With these assays, at 11 weeks, the second Wisconsin survey had a time-invariant adjustment factor of 1.102 (95% CrI: 0.836, 1.368) compared to a time-varying adjustment factor of 1.364 (95% CrI: 1.030, 1.712). For those assays that have a lower susceptibility to seroreversion the differences between time-invariant and time-varying adjustment factors tend to be substantially smaller.

We present unadjusted reported seroprevalence as well as the population-level estimate with time-invariant and time-varying adjustments in Table 2. Uncorrected estimates range from 0.9% in the Ohio survey to 20.8% in the New York City survey. The maximum difference we estimated between the uncorrected reported seroprevalence and the estimate with time-varying adjustments was 3.4 percentage points in the New York survey (20.8% versus 24.2%). Even with a long 17-week average period between infection and the survey, 2.7 percentage points of the difference were accounted for by the time-invariant adjustments due to the assay being one with a lower risk from seroreversion. However, the second-largest difference was 2.5 percentage points in the second Wisconsin sample (6.8% versus 9.3%). With shorter average time of 11 weeks post-infection but using an assay with higher risk from seroreversion, the time-invariant adjustments accounted for only 0.7 percentage points and the time-varying adjustments accounted for the remaining 1.8. In 11 of 15 surveys, the difference was less than two percentage points.

**Table 2:** Seroprevalence estimates. Adjusted estimates represent posterior medians and are accompanied by 95% credible intervals.

| Jurisdiction | Average Interval from Infection (weeks) | Risk from seroreversion | Reported | Time-invariant | Time-varying | Varying/Invariant |
| --- | --- | --- | --- | --- | --- | --- |
| United States | 18 | Low | 0.0524 | 0.063<br>(0.052, 0.092) | 0.068<br>(0.053, 0.116) | 1.063<br>(0.795, 1.690) |
| Arkansas |  |  |  |  |  |  |
| Phase 2 | 4 | Low | 0.0330 | 0.035<br>(0.014, 0.059) | 0.035<br>(0.014, 0.060) | 1.010<br>(0.376, 2.787) |
| Phase 5 | 9 | Low | 0.0620 | 0.069<br>(0.047, 0.098) | 0.070<br>(0.048, 0.103) | 1.018<br>(0.668, 1.588) |
| Phase 7 | 11 | Low | 0.1420 | 0.164<br>(0.117, 0.232) | 0.167<br>(0.119, 0.242) | 1.019<br>(0.672, 1.564) |
| California | 16 | Low | 0.0450 | 0.054<br>(0.037, 0.084) | 0.058<br>(0.038, 0.097) | 1.062<br>(0.653, 1.822) |
| Connecticut | 12 | Low | 0.0400 | 0.045<br>(0.026, 0.063) | 0.050<br>(0.029, 0.073) | 1.112<br>(0.605, 2.114) |
| Georgia | 17 | Low | 0.0860 | 0.104<br>(0.082, 0.153) | 0.111<br>(0.084, 0.186) | 1.062<br>(0.763, 1.676) |
| New York City | 18 | Low | 0.2076 | 0.235<br>(0.192, 0.282) | 0.242<br>(0.199, 0.290) | 1.031<br>(0.808, 1.320) |
| Ohio | 9 | High | 0.0090 | 0.009<br>(0.002, 0.018) | 0.011<br>(0.002, 0.021) | 1.154<br>(0.195, 6.310) |
| Puerto Rico | 17 | High | 0.0200 | 0.019<br>(0.009, 0.028) | 0.026<br>(0.012, 0.045) | 1.419<br>(0.711, 2.939) |
| Rhode Island | 4 | High | 0.0290 | 0.032<br>(0.018, 0.043) | 0.033<br>(0.020, 0.045) | 1.049<br>(0.592, 1.839) |
| Indiana |  |  |  |  |  |  |
| Phase 1 | 3 | High | 0.0109 | 0.012<br>(0.004, 0.016) | 0.012<br>(0.004, 0.016) | 1.036<br>(0.607, 1.774) |
| Phase 2 | 7 | High | 0.0150 | 0.016<br>(0.009, 0.021) | 0.018<br>(0.010, 0.024) | 1.097<br>(0.665, 1.800) |
| Wisconsin |  |  |  |  |  |  |
| Phase 1 | 10 | High | 0.0160 | 0.017<br>(0.007, 0.026) | 0.021<br>(0.008, 0.032) | 1.206<br>(0.508, 2.876) |
| Phase 2 | 11 | High | 0.0680 | 0.075<br>(0.057, 0.093) | 0.093<br>(0.070, 0.116) | 1.239<br>(0.887, 1.737) |

Arkansas data used the Beckman ACCESS assay. California and Georgia used BioRad Platelia. Connecticut used Vitros Ortho, S CoV2 IgG. New York City used DiaSorin. Ohio, Rhode Island, Indiana, and Wisconsin used Abbott Architect. Puerto Rico used Abbott Alinity i.

As a sensitivity analysis, we include studies from London including The Real-time Assessment of Commnity Transmission (REACT-2) study[34], the UK Office of National Statistics (ONS) COVID-19 infection survey[35], and the Biobank SARS-CoV-2 Serology Study[27]. Results from this analysis are available in Tables S1 and S2 as well as Figure S1 of the Supplementary Material. REACT-2 had a high risk of test-specific seroreversion and we note that the difference between the time-invariant and time-varying adjustments increases as the average interval from infection to increases. Conversely, the UK ONS COVID-19 infection survey and Biobank had a low risk of test-specific seroreversion. While the time-varying adjustment does result in a larger seroprevalence estimate which increases as the average interval from infection increases, it is not as large a difference as we observe in the REACT-2 study.

## Discussion

These results demonstrate that the effect of time-varying sensitivity is potentially large and that adjustment can reliably correct for it. Seroreversion decreases time-varying sensitivity more strongly for anti-nucleocapsid versus anti-spike assays, and when serological sampling occurs longer after infection [12]. Our seroreversion adjustment ratio, for instance, is higher for a Wisconsin study using an anti-nucleocapsid assay compared to an Indiana study using the same assay but with a 2-month shorter delay between infection and serological sampling. At 4 months post-infection time-varying sensitivity adjustment increased cumulative incidence compared to time-invariant adjustment by a factor of 1.03 for a New York City study using an anti-spike assay, but by 1.42 for a study in Puerto Rico using an anti-nucleocapsid assay. Seroreversion can therefore strongly bias cumulative incidence estimates. At its most, seroreversion increased estimated cumulative incidence 1.36-fold at several months after infection compared to the reported seroprevalence estimates. Assay-specific adjustments are needed to detect whether this bias is present. At the same time, the large relative adjustment for Puerto Rico corresponded to a small absolute difference, from 1.9% to 2.6% cumulative incidence. Seroreversion can lead to larger absolute differences later in a pandemic due to longer time since infection and greater cumulative incidence. Getting the adjustment right can be critical when comparing seroprevalence surveys done at different times relative to the peak infection period using different assays.

Our Bayesian seroreversion corrections yield more valid estimates of cumulative incidence as well as valid credible intervals. An advantage to Bayesian over frequentist approaches is that Bayesian models can incorporate uncertainty in test sensitivity and specificity, the rate of seroreversion, and estimates of seroprevalence through the use of priors. Because all of these are usually drawn from relatively small samples, they often have meaningful uncertainty. Prior COVIDVu seroprevalence analyses of California, Georgia, and the USA overall applied the anti-spike indirect ELISA adjustments of Shioda et al. to an anti-nucleocapsid direct ELISA. Time-varying sensitivity was also inferred from decreasing seroprevalence with time in a convenience sample, resulting in a 2-fold increase from reported seroprevalence to cumulative incidence adjusted for time-varying sensitivity [1,22,23,36]. But factors other than seroreversion could contribute to decreasing seroprevalence, such as convenience sampling becoming more or less representative of the general population with time [11,14,16]. We instead infer time-varying sensitivity from serological testing of prior-positives, yielding greater sensitivity and lower cumulative incidence estimates than reported in COVIDVu. Our time-varying sensitivity-adjusted cumulative incidence estimates are broadly consistent with two prior studies of the USA, including the results of Barber et al. that applied time-varying sensitivity adjustments accounting for only antibody isotype and antigen target for eight assays [26,37]. Our cumulative incidence estimates were higher than Barber et al. in Arkansas, likely due to the Arkansas seroprevalence study using a marketing database that overestimates seroprevalence in the general population[38,39].

This analysis improves upon surveys that estimated time-varying sensitivity using study-internal prior-positives [2,5,24,25]. These surveys assumed the same amount of seroreversion among prior-positives compared to other study participants, with the participants being representative of the general population. But prior-positives may differ from the general population with respect to attributes that can impact seroreversion, such as COVID-19 severity and age [14,40], though age may not strongly impact time-varying sensitivity for some assay designs [41,42]. Our model mitigates this selection bias by excluding prior-positive studies that sampled only particular groups, such as nursing home residents. We also assume the same amount of seroreversion among prior-positives compared to reported cases in the same location, instead of compared to the overall population. This approach also draws on prior-positive data from multiple studies, in contrast to more limited data from only one study population [12], and provides assay-specific time-varying sensitivity estimates for studies lacking data of prior-positives.

Despite these limitations, study-internal prior-positives usefully approximate time-varying sensitivity, especially in an outbreak lacking prior-positives data across multiple study populations. The study-internal proportion of prior-positives who tested seropositive better matches our time-varying sensitivity estimates than time-varying sensitivity estimates applied in three seroprevalence studies [1,22,23]. Our estimates still slightly exceeded this proportion of prior-positives, possibly due to lower sensitivity for the assay in question when compared to other anti-nucleocapsid direct ELISAs [12]. These results support other seroprevalence studies that inferred time-varying sensitivity-adjusted cumulative incidence using prior-positives within the study sample [2,25], including studies that relied on those cumulative incidence estimates to infer severity measures [5,24].

Our time-varying sensitivity adjustments provide more plausible cumulative incidence estimates for inferring severity measures. Seroreversion causes large relative changes in estimated cumulative incidence, inflating estimates of fatality and hospitalization rates from infection (IFR and IHR, respectively) [6,14,26]. This substantial inflation occurs even with small absolute differences in cumulative incidence estimates. Our analysis suggests IFR was overestimated for Connecticut, Indiana, and Rhode Island due to the absence of adjustments for time-varying sensitivity [43–45]. COVIDVu research applied time-varying sensitivity adjustments in analyses of California, Georgia, and the USA nationally [1,22,23]. Our analysis suggest these adjustments underestimated sensitivity and thereby underestimated IFR. Replacing COVIDVu cumulative incidence estimates with our estimates, for example, increased the national adult IFR estimate from 0.9% to 1.5%, closer to another published estimate [26]. Applying our time-varying sensitivity adjustments would also clarify whether seroreversion strongly [14] or mildly [6] biases an unusually high IFR estimate for Italy. Our results justify caution regarding IFR analyses that assume the same time-varying sensitivity trend across assay designs and antigen targets [6,19,20], given the impact of design and target on time-varying sensitivity. Multiple COVID-19 waves and underreporting of COVID-19 deaths [5,13,26,46] may also bias IFR estimates that use only an initial peak in reported deaths to infer time since infection [19,20], in contrast to the excess deaths used to infer time since infection in our approach.

Seroreversion adjustments for SARS-CoV-2 seroprevalence surveys may inform adjustments for surveys for other pathogens. The time-course of infections could be inferred from excess mortality, or by other surveillance methods for pathogens lacking weekly excess mortality estimates [47]. Design-specific adjustments could apply to assays targeting pathogens other than SARS-CoV-2 [48], though antigen-specific adjustments will likely vary. Future work could generate time-varying sensitivity corrections more applicable across pathogens by quantifying how antigen characteristics impact seroreversion. Seroreversion may differ, for example, for surface antigen epitopes exposed to soluble and membrane-bound immunoglobulins versus for epitopes hidden deeper within the pathogen, as with SARS-CoV-2 spike versus nucleocapsid protein, respectively.

Our approach provides an improved estimate of the cumulative incidence of COVID-19 at the time of survey. In virtually all circumstances, the uncorrected reported seroprevalence values underestimate our cumulative incidence estimates by 1.1-fold and often meaningfully more.

Such adjustments are critical for research and surveillance purposes requiring precise estimates of cumulative incidence, such as calculating infection fatality rates or discerning population-wide herd immunity levels. For other purposes, though, adjustments are less important. For example, few routine public health decisions during a pandemic turn on the difference between an unadjusted 4% and adjusted 6% cumulative incidence—absolute differences commonly observed in our data. Thus, simpler, less time-consuming “naive” methods that do not require waiting on the tabulation of mortality data for adjustments may suffice for some public health measures.

Several factors constrain our seroreversion analysis. We focus on serology studies performed before the introduction of SARS-CoV-2 vaccination to avoid detection of vaccine-induced antibodies and anti-spike vaccination reducing sensitivity of anti-nucleocapsid assays [49]. This approach, however, may be extended to post-vaccination time periods using seroreversion estimates for vaccine-induced antibodies. We choose to use excess mortality as an indicator of the lag between the average time of infection and the survey date because it was (a) readily available in a weekly time series and (b) less sensitive to temporal biases caused by shortages of tests, testing lags, and other factors that were common in 2020 than counts of cases or deaths.

However, this approach implicitly assumes IFR does not significantly vary with time when using excess mortality to infer time since infection, similar to other seroreversion analyses [14,19].

This assumption may not strongly bias inferred time when infections are concentrated in a narrow peak of deaths, or during time periods before factors such as vaccination and substantial transmission of variants impacted IFR [50,51]. Future work may test our methods’ robustness to changes in IFR. Further analysis may quantify how much time-varying sensitivity contributes to differences in reported seroprevalence within studies of the same population [52]. This would distinguish the impact of time-varying sensitivity from that of factors such as sampling bias [53].

Adjusting for seroreversion becomes more important with time during outbreaks. Our Bayesian time-varying sensitivity adjustments illustrate a promising method for accounting for antigen and assay characteristics in seroreversion corrections, providing more reliable estimates of time-varying cumulative incidence, post-infection immunity, and severity measures.

## Conclusions

Depending on the assay used, the naive use of tests with sensitivity that decays over time can seriously bias estimates of cumulative incidence of SARS-CoV-2 infection. The Bayesian approach assay-specific time-varying sensitivity adjustment developed in this paper can reliably correct for this bias. In seroprevalence studies conducted in the United States in 2020, adjusting for time-varying sensitivity increased cumulative incidence up to 1.4-fold. This implies that previous seroreversion corrections overestimated cumulative incidence in the United States up to 1.7-fold by not accounting for assay-specific effects. Similarly, estimates of the infection fatality rate can be substantially biased, with a corrected estimate in one study 1.7 times higher than the reported rate.

## Abbreviations

CI: confidence interval
COVID-19: coronavirus disease 2019
CrI: credible interval
ELISA: enzyme-linked immunosorbent assay
IFR: infection fatality rate
IHR: infection hospitalization rate
SARS-CoV-2: severe acute respiratory syndrome coronavirus 2

## Data availability

The data that support the findings of this study are openly available at https://data.mendeley.com/datasets/yccc76j326/1.

## Acknowledgments

The authors thank Dr. Karen A. Alroy for sharing additional seroprevalence data.

## References

1. Sullivan PS, et al. Severe Acute Respiratory Syndrome Coronavirus 2 Cumulative Incidence, United States, August 2020–December 2020. Clinical Infectious Diseases 2022; 74: 1141–1150.

2. Neuhauser H, et al. Nationally representative results on SARS-CoV-2 seroprevalence and testing in Germany at the end of 2020. Scientific Reports 2022; 12: 19492.

3. Stringhini S, et al. Seroprevalence of anti-SARS-CoV-2 IgG antibodies in Geneva, Switzerland (SEROCoV-POP): a population-based study. The Lancet 2020; 396: 313–319.

4. Lipsitch M, et al. Cross-reactive memory T cells and herd immunity to SARS-CoV-2. Nature Reviews Immunology 2020; 20: 709–713.

5. Levin AT, et al. Assessing the burden of COVID-19 in developing countries: systematic review, meta-analysis and public policy implications. BMJ Global Health 2022; 7: e008477.

6. Brazeau NF, et al. Estimating the COVID-19 infection fatality ratio accounting for seroreversion using statistical modelling. Communications Medicine 2022; 2: 54.

7. Buss LF, et al. Three-quarters attack rate of SARS-CoV-2 in the Brazilian Amazon during a largely unmitigated epidemic. Science 2021; 371: 288–292.

8. Meyer MJ, et al. Adjusting COVID-19 Seroprevalence Survey Results to Account for Test Sensitivity and Specificity. American Journal of Epidemiology 2022; 191: 681–688.

9. Müller SA, et al. Learning from serosurveillance for SARS-CoV-2 to inform pandemic preparedness and response. The Lancet 2023; 402: 356–358.

10. Hay JA, Routledge I, Takahashi S. Serodynamics: A primer and synthetic review of methods for epidemiological inference using serological data. Epidemics 2024; 49: 100806.

11. Kadelka S, et al. Correcting for Antibody Waning in Cumulative Incidence Estimation From Sequential Serosurveys. American Journal of Epidemiology 2024; 193: 777–786.

12. Owusu-Boaitey N, et al. Dynamics of SARS-CoV-2 seroassay sensitivity: a systematic review and modelling study. Eurosurveillance 2023; 28Published online: 25 May 2023.doi:10.2807/1560-7917.ES.2023.28.21.2200809.

13. Ward H, et al. Design and Implementation of a National Program to Monitor the Prevalence of SARS-CoV-2 IgG Antibodies in England Using Self-Testing: The REACT-2 Study. American Journal of Public Health 2023; 113: 1201–1209.

14. Takahashi S, et al. SARS-CoV-2 Serology Across Scales: A Framework for Unbiased Estimation of Cumulative Incidence Incorporating Antibody Kinetics and Epidemic Recency. American Journal of Epidemiology 2023; 192: 1562–1575.

15. Lohse S, et al. German federal-state-wide seroprevalence study of 1st SARS-CoV-2 pandemic wave shows importance of long-term antibody test performance. Communications Medicine 2022; 2: 52.

16. He D, et al. Resolving the enigma of Iquitos and Manaus: A modeling analysis of multiple COVID-19 epidemic waves in two Amazonian cities. Proceedings of the National Academy of Sciences 2023; 120: e2211422120.

17. Lalwani P, et al. SARS-CoV-2 seroprevalence and associated factors in Manaus, Brazil: baseline results from the DETECTCoV-19 cohort study. International Journal of Infectious Diseases 2021; 110: 141–150.

18. Haile SR, Kronthaler D. Potential for Bias in Prevalence Estimates when Not Accounting for Test Sensitivity and Specificity: A Systematic Review of COVID-19 Seroprevalence Studies. International Journal of Public Health 2025; 70: 1608343.

19. Pezzullo AM, et al. Age-stratified infection fatality rate of COVID-19 in the non-elderly population. Environmental Research 2023; 216: 114655.

20. Axfors C, Ioannidis JPA. Infection fatality rate of COVID-19 in community-dwelling elderly populations. European Journal of Epidemiology 2022; 37: 235–249.

21. Shioda K, et al. Estimating the Cumulative Incidence of SARS-CoV-2 Infection and the Infection Fatality Ratio in Light of Waning Antibodies. Epidemiology 2021; 32: 518–524.

22. Chamberlain AT, et al. Cumulative Incidence of SARS-CoV-2 Infections Among Adults in Georgia, United States, August to December 2020. The Journal of Infectious Diseases 2022; 225: 396–403.

23. Lamba K, et al. SARS-CoV-2 Cumulative Incidence and Period Seroprevalence: Results From a Statewide Population-Based Serosurvey in California. Open Forum Infectious Diseases 2021; 8: ofab379.

24. Mutevedzi PC, et al. Estimated SARS-CoV-2 infection rate and fatality risk in Gauteng Province, South Africa: a population-based seroepidemiological survey. International Journal of Epidemiology 2022; 51: 404–417.

25. Fantin R, et al. Estimating the cumulative incidence of SARS-CoV-2 infection in Costa Rica: modelling seroprevalence data in a population-based cohort. The Lancet Regional Health - Americas 2023; 27: 100616.

26. Barber RM, et al. Estimating global, regional, and national daily and cumulative infections with SARS-CoV-2 through Nov 14, 2021: a statistical analysis. The Lancet 2022; 399: 2351–2380.

27. Bešević J, et al. Persistence of SARS-CoV-2 antibodies over 18 months following infection: UK Biobank COVID-19 Serology Study. Journal of Epidemiology and Community Health 2024; 78: 105–108.

28. Office for National Statistics. Coronavirus (COVID-19) Infection Survey: methods and further information. 2023(https://www.ons.gov.uk/peoplepopulationandcommunity/healthandsocialcare/conditionsanddiseases/methodologies/covid19infectionsurveypilotmethodsandfurtherinformation).

29. Gelman A, Carpenter B. Bayesian Analysis of Tests with Unknown Specificity and Sensitivity. Journal of the Royal Statistical Society Series C: Applied Statistics 2020; 69: 1269–1283.

30. Centers for Disease Control and Prevention. Excess Deaths Associated with COVID-19. 2025(https://www.cdc.gov/nchs/nvss/vsrr/covid19/excess_deaths.htm). Accessed 8 September 2025.

31. Yadav AK, et al. Seroconversion among COVID-19 patients admitted in a dedicated COVID hospital: A longitudinal prospective study of 1000 patients. Medical Journal, Armed Forces India 2021; 77: S379–S384.

32. Zhou F, et al. Clinical course and risk factors for mortality of adult inpatients with COVID-19 in Wuhan, China: a retrospective cohort study. Lancet (London, England) 2020; 395: 1054–1062.

33. Rogan WJ, Gladen B. Estimating prevalence from the results of a screening test. American Journal of Epidemiology 1978; 107: 71–76.

34. Imperial College London. The REACT-2 programme. (https://www.imperial.ac.uk/medicine/research-and-impact/groups/react-study/studies/the-react-2-programme/). Accessed 29 July 2026.

35. Coronavirus (COVID-19) Infection Survey - Office for National Statistics. (https://www.ons.gov.uk/surveys/informationforhouseholdsandindividuals/householdandindividualsurveys/covid19infectionsurvey). Accessed 29 July 2026.

36. Shioda K, et al. Estimating the Cumulative Incidence of SARS-CoV-2 Infection and the Infection Fatality Ratio in Light of Waning Antibodies. Epidemiology 2021; 32: 518–524.

37. Irons NJ, Raftery AE. Estimating SARS-CoV-2 infections from deaths, confirmed cases, tests, and random surveys. Proceedings of the National Academy of Sciences 2021; 118: e2103272118.

38. Owusu-Boaitey N, et al. Agreement between seroprevalence- and model-based estimates of COVID-19 burden. Global Health Action 2026; 19: 2683286.

39. Cardenas VM, et al. State-wide random seroprevalence survey of SARS-CoV-2 past infection in a southern US State, 2020. PloS One 2022; 17: e0267322.

40. Renk H, et al. Robust and durable serological response following pediatric SARS-CoV-2 infection. Nature Communications 2022; 13: 128.

41. Einhauser S, et al. Time Trend in SARS-CoV-2 Seropositivity, Surveillance Detection- and Infection Fatality Ratio until Spring 2021 in the Tirschenreuth County—Results from a Population-Based Longitudinal Study in Germany. Viruses 2022; 14: 1168.

42. Radon K, et al. From first to second wave: follow-up of the prospective COVID-19 cohort (KoCo19) in Munich (Germany). BMC Infectious Diseases 2021; 21: 925.

43. Chan PA, et al. Seroprevalence of SARS-CoV-2 Antibodies in Rhode Island From a Statewide Random Sample. American Journal of Public Health 2021; 111: 700–703.

44. Mahajan S, et al. SARS-CoV-2 Infection Hospitalization Rate and Infection Fatality Rate Among the Non-Congregate Population in Connecticut. The American Journal of Medicine 2021; 134: 812–816.e2.

45. Levin AT, et al. Assessing the age specificity of infection fatality rates for COVID-19: systematic review, meta-analysis, and public policy implications. European Journal of Epidemiology 2020; 35: 1123–1138.

46. Owusu-Boaitey N, et al. Impact of cross-reactivity and herd immunity on SARS-CoV-2 pandemic severity. Infectious Diseases 2024; : 1–6.

47. Thompson WW, et al. Estimates of US influenza-associated deaths made using four different methods. Influenza and Other Respiratory Viruses 2009; 3: 37–49.

48. Yao M, et al. Surveillance of Plasmodium vivax transmission using serological models in the border areas of China–Myanmar. Malaria Journal 2022; 21: 69.

49. Dhakal S, et al. Reconsideration of Antinucleocapsid IgG Antibody as a Marker of SARS-CoV-2 Infection Postvaccination for Mild COVID-19 Patients. Open Forum Infectious Diseases 2023; 10: ofac677.

50. De Boer PT, et al. Age-specific severity of severe acute respiratory syndrome coronavirus 2 in February 2020 to June 2021 in the Netherlands. Influenza and Other Respiratory Viruses 2023; 17: e13174.

51. Perez-Guzman PN, et al. Epidemiological drivers of transmissibility and severity of SARS-CoV-2 in England. Nature Communications 2023; 14: 4279.

52. Murhekar MV, et al. SARS-CoV-2 seroprevalence among the general population and healthcare workers in India, December 2020–January 2021. International Journal of Infectious Diseases 2021; 108: 145–155.

53. Bartig S, et al. Corona Monitoring Nationwide (RKI-SOEP-2): Seroepidemiological Study on the Spread of SARS-CoV-2 Across Germany. Jahrbücher für Nationalökonomie und Statistik 2023; 243: 431–449.

