## Supplemental for "Estimating COVID-19 Cumulative Incidence from Seroprevalence Surveys accounting for Time-Varying Seroreversion: A Fully Bayesian Methodology"

**Web Material**

**Table of contents**

Web Appendix 1. Search procedure and selection criteria for seroprevalence surveys

Web Appendix 2. List of seroprevalences surveys and risk of bias assessment

Web Appendix 3. References

Web Appendix 4: Supplementary Results for the London Seroprevalence Surveys

**Web Appendix 1. Search procedure and selection criteria for seroprevalence surveys**

A literature search for seroprevalence surveys was performed weekly until May 30, 2025 using previously published methods (1). Surveys meeting any of the following criteria were excluded:

-	Sample more than 2 weeks after the onset of widespread SARS-CoV-2 vaccination in the sampled population, without confirmation from another source that the vaccination rate was negligible (2)

-	Sample only a population outside of the USA or Puerto Rico

-	Include only convenience samples such as residual sera from clinical testing (3–5)

-	Report gender ratios with <35% of a reported gender and no cited evidence that this gender ratio matched the sampled population

-	Report only seroprevalence adjusted for sensitivity and specificity, with no method for removing this adjustment

-	Use only the Wondfo serological assay (6)

-	Report only results following serial or parallel testing with multiple assays, instead of results from one assay

Only studies meeting each of the following criteria were included:

-	Sample a population within the USA or Puerto Rico

-	Estimate seroprevalence in a representative sample, i.e. random selection from a sample frame representative of the general population or sampling a majority of the total population (7–9)

-	Available in English or in a form readily translatable to English

-	Report median week of sampling, or sampling start-week and end-week

-	Report the number of individuals tested

-	Provide sufficient information to identify the assay used, or to ascertain assay sensitivity and specificity with confidence intervals

-	Sample a location with weekly excess deaths data available from March 1, 2020 to 21 days after the median week of serological sampling, with reported deaths used for calculating excess mortality instead of using modeled inferences from other locations (10,11)

These criteria led to the inclusion of ten surveys of USA states, New York City, and Puerto Rico ((12–23), correspondence with Dr. Karen A. Alroy). A national USA seroprevalence study was also included whose sample overlapped with state-wide surveys of California and Georgia (13,15,24). Web Appendix 2 provides a list of included and excluded surveys.

**Web Appendix 2. List of seroprevalences surveys**

The following repository contains a list of excluded and included seroprevalence studies, along with a risk of bias assessment with respect to response rate (8). They can be accessed at <https://data.mendeley.com/datasets/yccc76j326/1>.

**Web Appendix 3. References**

1.	Levin AT, Owusu-Boaitey N, Pugh S, et al. Assessing the burden of COVID-19 in developing countries: systematic review, meta-analysis and public policy implications. *BMJ Global Health*. 2022;7(5):e008477.

2.	Dhakal S, Yu T, Yin A, et al. Reconsideration of Antinucleocapsid IgG Antibody as a Marker of SARS-CoV-2 Infection Postvaccination for Mild COVID-19 Patients. *Open Forum Infect Dis*. 2022;10(1):ofac677.

3.	Boyce RM, Shook-Sa BE, Aiello AE. A Tale of 2 Studies: Study Design and Our Understanding of Severe Acute Respiratory Syndrome Coronavirus 2 Seroprevalence. *Clin Infect Dis*. 2020;73(9):e3124–e3126.

4.	Byambasuren O, Dobler CC, Bell K, et al. Comparison of seroprevalence of SARS-CoV-2 infections with cumulative and imputed COVID-19 cases: Systematic review. *PLOS ONE*. 2021;16(4):e0248946.

5.	Gajda M, Kowalska M, Zejda JE. Impact of Two Different Recruitment Procedures (Random vs. Volunteer Selection) on the Results of Seroepidemiological Study (SARS-CoV-2). *International Journal of Environmental Research and Public Health*. 2021;18(18):9928.

6.	Silveira MF, Mesenburg MA, Dellagostin OA, et al. Time-dependent decay of detectable antibodies against SARS-CoV-2: A comparison of ELISA with two batches of a lateral-flow test. *Braz J Infect Dis*. 2021;25(4):101601.

7.	Current Trends Pilot Study of a Household Survey to Determine HIV Seroprevalence. 1991;(<https://www.cdc.gov/mmwr/preview/mmwrhtml/00001871.htm>). (Accessed October 31, 2024)

8.	Bobrovitz N, Noël K, Li Z, et al. SeroTracker-RoB: A decision rule-based algorithm for reproducible risk of bias assessment of seroprevalence studies. *Research Synthesis Methods*. 2023;14(3):414–426.

9.	Centers for Disease Control and Prevention. Overview of CASPER. *CASPER*. 2024;(<https://www.cdc.gov/casper/php/overview/index.html>)

10.	[Karlinsky A, Kobak D. Tracking excess mortality across countries during the COVID-19 pandemic with the World Mortality Dataset. *Elife* [electronic article]. 2021;10. (](http://paperpile.com/b/jXBeYu/Dok8G)<http://dx.doi.org/10.7554/eLife.69336>)

11.	Msemburi W, Karlinsky A, Knutson V, et al. The WHO estimates of excess mortality associated with the COVID-19 pandemic. *Nature*. 2023;613(7942):130–137.

12.	Cardenas VM, Kennedy JL, Williams M, et al. State-wide random seroprevalence survey of SARS-CoV-2 past infection in a southern US State, 2020. *PLOS ONE*. 2022;17(4):e0267322.

13.	Lamba K, Bradley H, Shioda K, et al. SARS-CoV-2 Cumulative Incidence and Period Seroprevalence: Results From a Statewide Population-Based Serosurvey in California. *Open Forum Infect Dis*. 2021;8(8):ofab379.

14.	Mahajan S, Srinivasan R, Redlich CA, et al. Seroprevalence of SARS-CoV-2-Specific IgG Antibodies Among Adults Living in Connecticut: Post-Infection Prevalence (PIP) Study. *Am J Med*. 2021;134(4):526–534.e11.

15.	Chamberlain AT, Toomey KE, Bradley H, et al. Cumulative Incidence of SARS-CoV-2 Infections Among Adults in Georgia, United States, August to December 2020. *J Infect Dis*. 2022;225(3):396–403.

16.	[Kline D, Li Z, Chu Y, et al. Estimating seroprevalence of SARS-CoV-2 in Ohio: A Bayesian multilevel poststratification approach with multiple diagnostic tests. *Proc Natl Acad Sci U S A* [electronic article]. 2021;118(26). (](http://paperpile.com/b/jXBeYu/lq9J1)<http://dx.doi.org/10.1073/pnas.2023947118>)

17.	Chan PA, King E, Xu Y, et al. Seroprevalence of SARS-CoV-2 Antibodies in Rhode Island From a Statewide Random Sample. 2021;(<https://ajph.aphapublications.org/doi/10.2105/AJPH.2020.306115>). (Accessed October 31, 2024)

18.	Menachemi N, Yiannoutsos CT, Dixon BE, et al. Population Point Prevalence of SARS-CoV-2 Infection Based on a Statewide Random Sample - Indiana, April 25-29, 2020. *MMWR Morb Mortal Wkly Rep*. 2020;69(29):960–964.

19.	Zeek A. IUPUI, ISDH release findings from Phase 2 of COVID-19 testing in Indiana. *News at IU*. 2020;(<https://news.iu.edu/live/news/26933-iupui-isdh-release-findings-from-phase-2-of>)

20.	Duszynski TJ, Fadel W, Wools-Kaloustian KK, et al. Association of Health Status and Nicotine Consumption with SARS-CoV-2 positivity rates. *BMC Public Health*. 2021;21(1):1786.

21.	Malecki K, Nikodemova M, Schultz AA, et al. Population changes in seroprevalence among a statewide sample in the United States. 2020;(<http://medrxiv.org/lookup/doi/10.1101/2020.12.18.20248479>)

22.	Parrott JC, Maleki AN, Vassor VE, et al. Prevalence of SARS-CoV-2 Antibodies in New York City Adults, June–October 2020: A Population-Based Survey. *J Infect Dis*. 2021;224(2):188–195.

23.	Puerto Rico Department of Health. A rapid community initial assessment to estimate the seroprevalence of COVID-19. Puerto Rico Science, Technology, and Research Trust; 2020.(<https://sites.google.com/view/casperpr/an%C3%A1lisis-y-resultados>)

24.	Sullivan PS, Siegler AJ, Shioda K, et al. Severe Acute Respiratory Syndrome Coronavirus 2 Cumulative Incidence, United States, August 2020–December 2020. *Clin Infect Dis*. 2021;74(7):1141–1150.

**Web Appendix 4: Supplementary Results for the London Seroprevalence Surveys**

**Table S1**: Posterior estimates of the adjustment factor, $B$, comparing reported seroprevalence to adjusted under both time invariant and time-varying adjustments for the London sensitivity analysis. Estimates represent posterior medians and are accompanied by 95% credible intervals.

| Study | Average Interval from Infection (weeks) | Risk from seroreversion | Time-invariant/Reported | | Time-varying/Reported | |
| --- | --- | --- | --- | --- | --- | --- |
|  |  |  | Est.  (95% CrI) | $P(B>1)$ | Est.  (95% CrI) | $P(B>1)$ |
| REACT-2 |  |  |  |  |  |  |
| Phase 1 | 14 | High | 1.140  (1.034, 1.335) | 0.997 | 1.598  (1.250, 2.418) | 1.000 |
| Phase 2 | 19 | High | 1.140  (1.027, 1.337) | 0.993 | 1.988  (1.373, 3.611) | 1.000 |
| Phase 3 | 26 | High | 1.140  (1.036, 1.335) | 0.998 | 3.020  (1.645, 7.472) | 1.000 |
| Phase 4 | 31 | High | 1.140  (1.034, 1.334) | 0.997 | 4.332  (1.923, 11.62) | 1.000 |
| ONS |  |  |  |  |  |  |
| Phase 1 | 9 | Low | 1.094  (0.554, 1.640) | 0.636 | 1.118  (0.558, 1.673) | 0.668 |
| Phase 2 | 13 | Low | 1.108  (0.727, 1.498) | 0.722 | 1.149  (0.750, 1.555) | 0.772 |
| Phase 3 | 17 | Low | 1.092  (0.665, 1.508) | 0.665 | 1.140  (0.691, 1.583) | 0.735 |
| Phase 4 | 21 | Low | 1.091  (0.776, 1.373) | 0.723 | 1.155  (0.817, 1.483) | 0.827 |
| Phase 5 | 25 | Low | 1.093  (0.857, 1.287) | 0.810 | 1.175  (0.917, 1.452) | 0.918 |
| Phase 6 | 29 | Low | 1.094  (0.898, 1.250) | 0.865 | 1.194  (0.953, 1.473) | 0.955 |
| Phase 7 | 31 | Low | 1.098  (0.926, 1.227) | 0.907 | 1.211  (0.995, 1.512) | 0.973 |
| Phase 8 | 33 | Low | 1.099  (0.961, 1.201) | 0.949 | 1.223  (1.028, 1.565) | 0.986 |
| Biobank |  |  |  |  |  |  |
| Phase 2 | 12 | Low | 1.095  (0.903, 1.198) | 0.922 | 1.130  (0.932, 1.250) | 0.950 |
| Phase 3 | 18 | Low | 1.098  (0.933, 1.193) | 0.940 | 1.151  (0.970, 1.289) | 0.965 |
| Phase 4 | 29 | Low | 1.097  (0.942, 1.190) | 0.945 | 1.195  (1.010, 1.440) | 0.978 |
| The REACT-2 study used the Fortress Diagnostic assay. The ONS and Biobank studies used the University of Oxford assay. | | | | | | |

**Table S2**: Seroprevalence estimates for the London sensitivity analysis. Adjusted estimates represent posterior medians and are accompanied by 95% credible intervals.

| Study | Average Interval from Infection (weeks) | Risk from seroreversion | Median  date | Reported | Time-invariant | Time-varying | Varying/  Invariant |
| --- | --- | --- | --- | --- | --- | --- | --- |
| REACT-2 |  |  |  |  |  |  |  |
| Phase 1 | 14 | High | July 1 | 0.109 | 0.124  (0.113, 0.145) | 0.174  (0.136, 0.263) | 1.399  (1.131, 1.953) |
| Phase 2 | 19 | High | Aug 6 | 0.079 | 0.090  (0.081, 0.106) | 0.157  (0.108, 0.285) | 1.739  (1.230, 2.999) |
| Phase 3 | 26 | High | Sept 21 | 0.080 | 0.091  (0.082, 0.106) | 0.240  (0.131, 0.594) | 2.634  (1.459, 6.411) |
| Phase 4 | 31 | High | Nov 3 | 0.080 | 0.091  (0.083, 0.107) | 0.347  (0.154, 0.932) | 3.764  (1.700, 9.927) |
| ONS |  |  |  |  |  |  |  |
| Phase 1 | 9 | Low | May 25 | 0.150 | 0.164  (0.083, 0.246) | 0.168  (0.084, 0.251) | 1.025  (0.475, 2.181) |
| Phase 2 | 13 | Low | June 22 | 0.154 | 0.171  (0.112, 0.231) | 0.177  (0.116, 0.240) | 1.035  (0.625, 1.696) |
| Phase 3 | 17 | Low | July 20 | 0.113 | 0.123  (0.075, 0.170) | 0.129  (0.078, 0.179) | 1.048  (0.598, 1.846) |
| Phase 4 | 21 | Low | Aug 17 | 0.103 | 0.112  (0.080, 0.141) | 0.119  (0.084, 0.153) | 1.057  (0.726, 1.552) |
| Phase 5 | 25 | Low | Sept 14 | 0.115 | 0.126  (0.098, 0.148) | 0.135  (0.106, 0.167) | 1.080  (0.826, 1.429) |
| Phase 6 | 29 | Low | Oct 12 | 0.111 | 0.121  (0.100, 0.139) | 0.132  (0.106, 0.164) | 1.096  (0.882, 1.395) |
| Phase 7 | 31 | Low | Nov 4 | 0.127 | 0.139  (0.118, 0.156) | 0.154  (0.126, 0.192) | 1.108  (0.909, 1.411) |
| Phase 8 | 33 | Low | Dec 7 | 0.160 | 0.176  (0.154, 0.192) | 0.196  (0.164, 0.250) | 1.118  (0.941, 1.433) |
| Biobank |  |  |  |  |  |  |  |
| Phase 2 | 12 | Low | June 16 | 0.104 | 0.114  (0.094, 0.125) | 0.117  (0.097, 0.130) | 1.033  (0.915, 1.170) |
| Phase 3 | 18 | Low | July 26 | 0.116 | 0.127  (0.108, 0.138) | 0.134  (0.112, 0.150) | 1.050  (0.930, 1.194) |
| Phase 4 | 29 | Low | Oct 9 | 0.124 | 0.136  (0.117, 0.148) | 0.148  (0.125, 0.179) | 1.093  (0.950, 1.324) |
| The REACT-2 study used the Fortress Diagnostic assay. The ONS and Biobank studies used the University of Oxford assay. | | | | | | | |

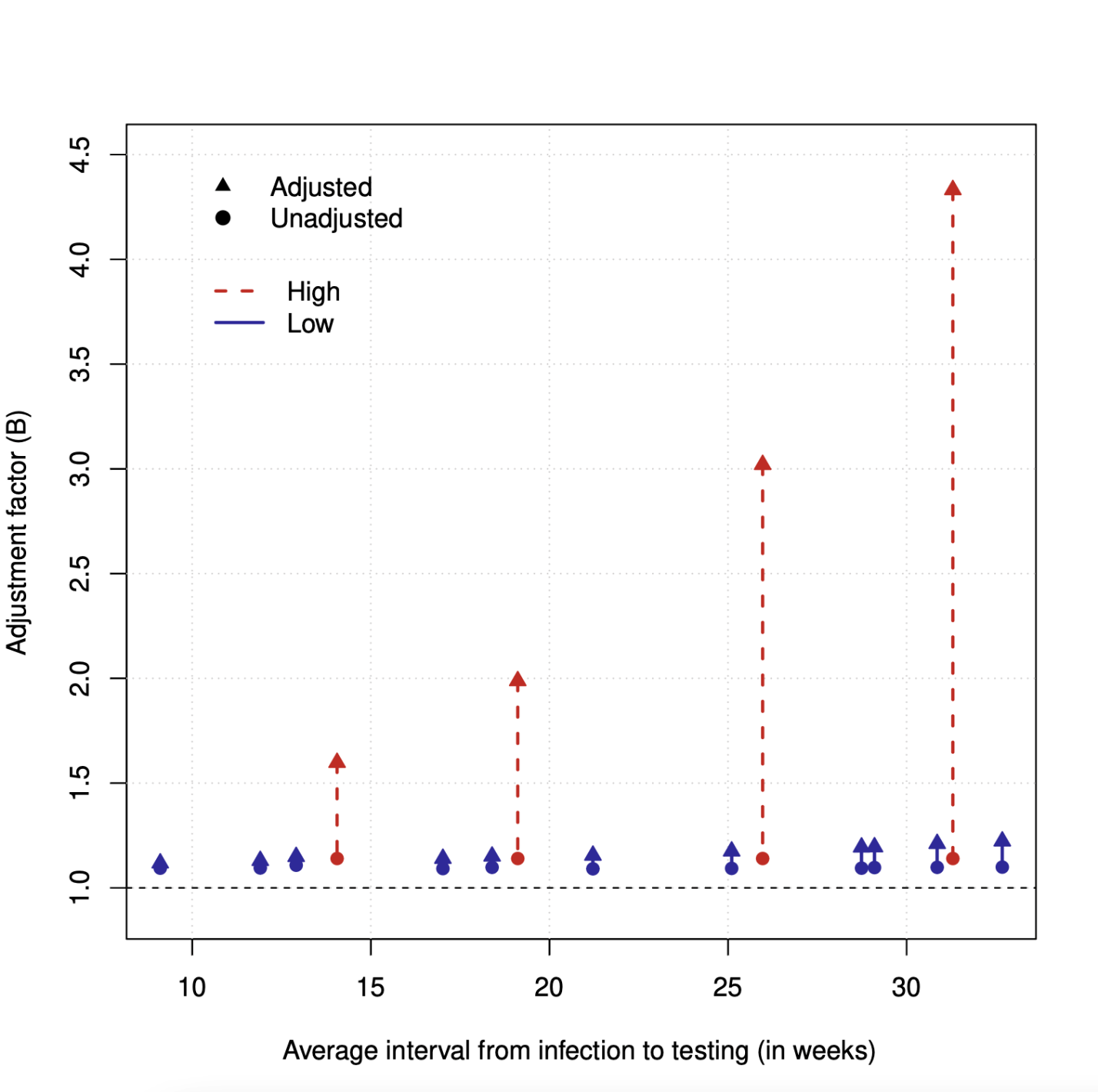

**Figure S1**: Adjustment factor $B$ for the London sensitivity analysis when calculated using the time-invariant sensitivity (circles) versus calculated using time-varying sensitivity (triangles). Solid blue indicates low test-specific seroreversion risk while dashed red indicates high seroreversion risk.
